# Acute kidney injury at early stage as a negative prognostic indicator of patients with COVID-19: a hospital-based retrospective analysis

**DOI:** 10.1101/2020.03.24.20042408

**Authors:** Shen Xu, Lin Fu, Jun Fei, Hui-Xian Xiang, Ying Xiang, Zhu-Xia Tan, Meng-Die Li, Fang-Fang Liu, Ying Li, Ming-Feng Han, Xiu-Yong Li, De-Xin Yu, Hui Zhao, De-Xiang Xu

## Abstract

Coronavirus disease 2019 (COVID-19) is a newly emerged infection of severe acute respiratory syndrome coronavirus-2 (SARS-CoV-2) and has been pandemic all over the world. This study described acute kidney injury (AKI) at early stage of COVID-19 and its clinical significance. Three-hundred and fifty-five COVID-19 patients with were recruited and clinical data were collected from electronic medical records. Patient’s prognosis was tracked and risk factors of AKI was analyzed. Of 355 COVID-19 patients, common, severe and critical ill cases accounted for 63.1%, 16.9% and 20.0%, respectively. On admission, 56 (15.8%) patients were with AKI. Although AKI was more common in critical ill patients with COVID-19, there was no significant association between oxygenation index and renal functional indices among COVID-19 patients with AKI. By multivariate logistic regression, male, older age and comorbidity with diabetes were three important independent risk factors predicting AKI among COVID-19 patients. Among 56 COVID-19 patients with AKI, 33.9% were died on mean 10.9 day after hospitalization. Fatality rate was obviously higher among COVID-+19 patients with AKI than those without AKI (*RR*=7.08, *P*<0.001). In conclusion, male elderly COVID-19 patients with diabetes are more susceptible to AKI. AKI at early stage may be a negative prognostic indicator for COVID-19.

## 1 Introduction

Since December 2019, a cluster of acute respiratory illness patients with unclear causes were found in several hospitals in Wuhan city, Hubei province, China (WHO. Novel coronavirus China. 2020; https://www.who.int/csr/don/12-january-2020-novel-coronavirus-china/en/). On February12, 2020, the International Committee on Taxonomy of Viruses declared the official classification of this novel coronavirus as severe acute respiratory syndrome coronavirus 2 (SARS-CoV-2). The World Health Organization (WHO) pronounced the official name of SARS-CoV-2-induced disease as coronavirus disease 2019 (COVID-19). Since the first patient was reported in December 2019 in Wuhan city, Hubei province, China, COVID-19 broke out quickly and had been pandemic all over the world ^1^. Until 15th March, 2020, more than 150,000 cases were confirmed to be infected with SARS-CoV-2 and more than 5000 cases died from SARS-CoV-2 infection all over the world (China CDC Weekly. Novel coronavirus China. 2020; http://weekly.chinacdc.cn/index.htm).

SARS-CoV-2 is mainly transmitted by droplets or direct contact and infected through respiratory tract ^2^. In addition, SARS-CoV-2 can also be transmitted by feces^3^. Accumulating data demonstrate that SARS-CoV-2 uses angiotensin-converting enzyme 2 (ACE2) to enter human cells, mainly pulmonary epithelial cells, and probably cardiomyocytes and renal tubular epithelial cells ^4-6^. The main symptoms and signs of COVID-19 patients are fever, companied by dry cough, dyspnea, diarrhea, fatigue and lymphopenia ^7-11^. Although only a few cases died in mild COVID-19 patients, death risk was rapidly increased among critical ill patients with the fatality rate even more than 50% ^12^. A large number of clinical data have revealed that infection with SARS-CoV-2 not only causes severe acute respiratory syndrome but also multiple organ injuries, including myocardial dysfunction, hepatic injury and even acute renal failure (AKI) ^13^. Nevertheless, what factors influence the multiple organ injuries, especially AKI, during the pathogenesis of COVID-19 remains unclear. Moreover, the clinical significance of AKI needs to be further clarified.

The objective of this study is to describe the clinical and laboratory features of AKI at early stage of 355 COVID-19 patients in two hospitals from different regions. We showed that male elderly COVID-19 patients with diabetes mellitus were more susceptible to AKI. We provide evidence that the development of AKI at early stage may be a potential negative prognostic indicator for survival of COVID-19 patients.

## 2 Methods

### 2.1 Study design and participants

In the present study, 200 patients, who were diagnosed as COVID-19 from January 1 to January 30, 2020, were recruited from Union Hospital of Huazhong University of Science and Technology, in Wuhan city, Hubei Province, China. One-hundred and fifty-five patients, who were diagnosed as COVID-19 from January 25 to February 20, 2020, were recruited from the Second People’s Hospital of Fuyang City, in Anhui province, China. Union Hospital of Huazhong University of Science and Technology is one of COVID-19-designated hospitals in Wuhan City. The Second Affiliated Hospital of Anhui Medical University sent a medical team to this hospital to recuse COVID-19 patients. The Second People’s Hospital of Fuyang City is one of COVID-19-designated hospitals in Anhui province. The Second Affiliated Hospital of Anhui Medical University sent a specialist group to guide COVID-19 treatment. All COVID-19 patients were clinically diagnosed on basis of typical clinical manifestations accompanied with characteristic chest radiology changes. Real-time RT-PCR was used to detect SARS-CoV-2 RNA and confirm infection with SARS-CoV-2. The patient with negative result on SARS-CoV-2 RNA detection was excluded from this study. All COVID-19 patients were eligible in this study. Each COVID-19 patient gave advanced oral consent. The present study was approved by the Ethics Committee of Anhui Medical University.

### 2.2 Data collection

The medical record of each COVID-19 patient was evaluated. Following data were collected from the electronic medical records: demographic information, preexisting comorbidities, including chronic obstructive pulmonary disease, hepatic disease, cardiovascular disease, hypertension, diabetes and other disease. Patient’s signs and symptoms, chest computed tomographic (CT) scan and laboratory test results were also collected. The dates of onset, admission and death were recorded. The onset time was defined as the date when patients’ any symptom and sign were found.

### 2.3 Laboratory testing

Patient’s pharyngeal swab specimens were collected for extraction of SARS-CoV-2 RNA. Real-time RT-PCR was used to detect viral nucleic acid using a COVID-19 nucleic acid detection kit following experimental instructions (Shanghai bio-germ Medical Technology Co Ltd). Viral RNA extraction and nucleic acid detection were executed by either the Center for Disease Control and Prevention of Wuhan City or the Center for Disease Control and Prevention of Fuyang City. Other laboratory tests, such as routine blood test, hepatic function, creatinine, urea nitrogen, uric acid, creatine kinase (CK) and oxygenation index were examined on admission. All laboratory tests were analyzed by the clinical laboratory of either the Union Hospital of Huazhong University of Science and Technology or the Second People’s Hospital of Fuyang City.

### 2.4 Statistical analysis

All statistical analyses were performed using SPSS 22.0 software. Categorical variables were expressed with frequencies and percentages. Continuous variables were shown using median and mean values. Means for continuous variables were compared with independent-samples *t* tests when the data were normally distributed; if not, the Mann-Whitney test was used. Proportions for categorical variables were compared with the *chi-square* and *Fisher’s* exact test. Univariable logistic regression between basic disease or different parameter and demise was performed. Moreover, the main risks related with demise were examined using multivariable logistic regression models adjusted for potential confounders. Statistical significance was determined at *P*<0.05.

## 3 Results

### 3.1 Demographic and clinical characteristics of COVID-19 patients

The clinical characteristics of 355 COVID-19 patients were analyzed. The most common symptom of COVID-19 patients was fever (75.0%), followed by cough (48.5%), diarrhea (36.6%) and fatigue (32.1%). Of 355 patients, common cases, defined as oxygenation index higher than 300, accounted for 63.1% (224/355). Severe cases, whose oxygenation index was from 200 to 300, accounted for 16.9% (60/355). Critical ill cases, oxygenation index lower than 200, accounted for another 20.0% (71/355). Moreover, 50.1% COVID-19 patients had at least one of comorbidity with either diabetes (41.4%) or hypertension (35.2%). The blood routine indices were analyzed. As shown in Table 1, mean WBC count was 5.67×10^9^/L. In addition, mean neutrophil and lymphocyte counts was 4.21×10^9^/L and 1.02×10^9^/L, respectively (Table 1). As shown in Table, 38% COVID-19 patients have neutrophil percentage above normal range. By contrast, 46.8% COVID-19 patients have lymphocyte percentage below normal range. In addition, 33% COVID-19 patients have hemoglobin below normal range (Table 1). Hepatic functional indices were then analyzed. As shown in Table 1, the levels of serum total bilirubin, direct bilirubin, total protein and albumin were 14.16 μmol/L, 5.17 μmol/L, 67.30 g/L and 38.53 g/L, respectively. Moreover, serum ALT and AST were 35.03 and 40.76 U/L, respectively. As shown in Table 1, 25.6% COVID-19 patients have ALT above normal range. In addition, 28.7% COVID-19 patients have AST above normal range. Next, myocardial functional indices are shown in Table 1. Mean creatine kinase, LDH and D-dimer were 257.69 U/L, 296.42 U/L and 2762.41 μg/L, respectively. As shown in Table 1, 45.6% COVID-19 patients have LDH above normal range. In addition, 71.8% COVID-19 patients have D-dimer above normal range.

**Table 1.** Clinical characteristics among 355 COVID-19 patients.

|  | Mean (Min, Max) | Above<br>Normal (%) | Below<br>Normal (%) |
| --- | --- | --- | --- |
| **Blood routine indices** |  |  |  |
| White Blood Cell (10<sup>9</sup>/L) | 5.67 (1.67, 28.25) | 20 (5.6) | 103 (29.0) |
| Neutrophil (10<sup>9</sup>/L) | 4.21 (0.79, 24.91) | 45 (12.7) | 26 (7.3) |
| Neutrophil Percent (%) | 70.95 (39.10, 95.80) | 135 (38.0) | 1 (0.3) |
| Lymphocyte (10<sup>9</sup>/L) | 1.02 (0.13, 2.69) | 0 (0) | 120 (33.8) |
| Lymphocyte Percent (%) | 20.92 (1.08, 50.10) | 12 (3.4) | 166 (46.8) |
| Monocytes (10<sup>9</sup>/L) | 0.38 (0.00, 1.58) | 9 (2.5) | 17 (4.8) |
| Red Blood Cell (10<sup>12</sup>/L) | 4.26 (1.86, 5.80) | 8 (2.3) | 58 (16.3) |
| Hemoglobin (g/L) | 131.44 (51.00, 171.10) | — | 117 (33.0) |
| **Liver functional indices** |  |  |  |
| Total Bilirubin (μmol/L) | 14.16 (0.70, 511.60) | 66 (18.6) | 2 (0.6) |
| Direct Bilirubin (μmol/L) | 5.17 (0.50, 252.70) | 38 (10.7) | — |
| Total Protein (g/L) | 67.30 (44.10, 90.00) | 22 (6.2) | 68 (19.2) |
| Albumin (g/L) | 38.53 (18.20, 56.10) | 5 (1.4) | 91 (25.6) |
| ALT (U/L) | 35.03 (1.00, 414.00) | 91 (25.6) | — |
| AST (U/L) | 40.76 (10.00, 475.00) | 102 (28.7) | — |
| LDL (mmol/L) | 1.79 (0.29, 4.78) | 24 (6.8) | — |
| HDL (mmol/L) | 1.14 (0.32, 7.27) | 22 (6.2) | 58 (16.3) |
| **Renal functional indices** |  |  |  |
| Creatinine (μmol/L) | 72.42 (32.00, 355.00) | 26 (7.3) | 83 (23.4) |
| Urea nitrogen (mmol/L) | 4.91 (1.10, 37.50) | 42 (11.8) | 49 (13.8) |
| Uric acid (μmol/L) | 259.68 (29.00, 677.00) | 26 (7.3) | 4 (1.1) |
| **Myocardial functional indices** |  |  |  |
| Creatine Kinase (U/L) | 257.69 (16, 30339) | 51 (14.4) | 1 (0.3) |
| LDH (U/L) | 296.42 (7, 1801) | 162 (45.6) | — |
| D-Dimer (μg/L) | 2762.41 (10, 382000) | 255 (71.8) | — |
| **Blood Electrolytes** |  |  |  |
| K | 3.85 (2.19, 6.19) | 5 (1.4) | 76 (21.4) |
| Na | 137.92 (113.00, 148.00) | 8 (2.3) | 55 (15.5) |
| Cl | 101.47 (76.00, 116.00) | 70 (19.7) | 20 (5.6) |
| Ca | 2.22 (1.68, 2.73) | 6 (1.7) | 194 (54.6) |
| P | 1.04 (0.38, 1.97) | 10 (2.8) | 139 (39.2) |
| Mg | 0.86 (0.57, 1.30) | 2 (0.6) | 85 (23.9) |

### 3.2 AKI is companied at the early stage of COVID-19 pneumonia

Renal functional indices, as determined by serum creatinine, urea nitrogen and uric acid, were measured among 355 COVID-19 patients. As shown in Table 1, mean urea nitrogen was 4.9±3.3 mmol/L. The levels of serum creatinine and uric acid were 72.4 ±34.7 and 258.9±102.8 μmol/L, respectively. Blood electrolytes are presented in Table 1. The concentrations of blood K, Na, Cl, Ca, P and Mg were 3.85, 137.92, 101.47, 2.22, 1.04 and 0.86 μmol/L, respectively. The association between AKI and the severity of COVID-19 was analyzed. As shown in Table 2, the levels of serum creatinine, urea nitrogen and uric acid were higher in critical ill patients than those of mild patients. In addition, the levels of serum urea nitrogen were higher in severe patients than those of mild patients (Table 2). The correlationship between oxygenation index and renal functional indices was then analyzed. There was no significant association between oxygenation index and all renal functional indices among COVID-19 patients with AKI (Fig.1A-C). As shown in Fig. 1, a negative correlation was observed between oxygenation index and blood urea nitrogen among COVID-19 patients without AKI. There was no significant association between oxygenation index and creatinine as well as uric acid among COVID-19 patients without AKI (Fig.1D-E). AKI was defined as any of renal functional indices beyond normal range ^14^. Among 355 COVID-19 patients, 56 (15.8%) patients were with AKI on admission (Table 2). Although 24 cases were with AKI among mild COVID-19 patients, AKI is more common in critical ill patients with COVID-19 (Table 2, 10.7% in mild cases vs 18.3% in severe cases vs 29.2% in critical ill cases, *P*<0.01).

**Table 2.**
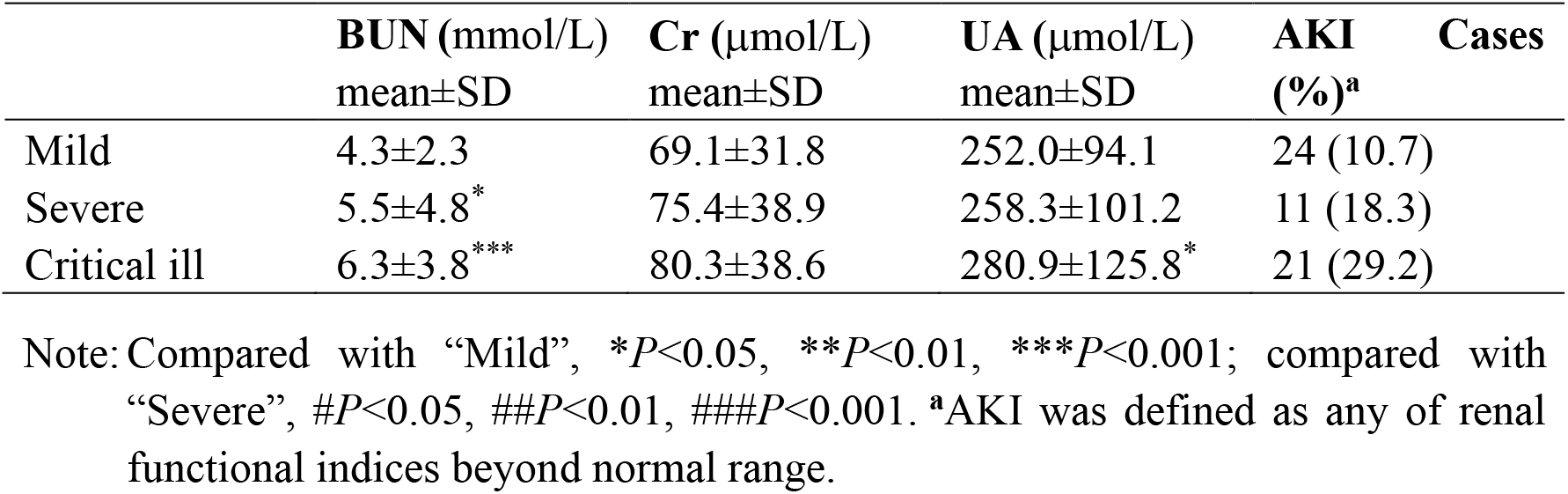
The association between the severity and renal function indices.

**Fig. 1.**
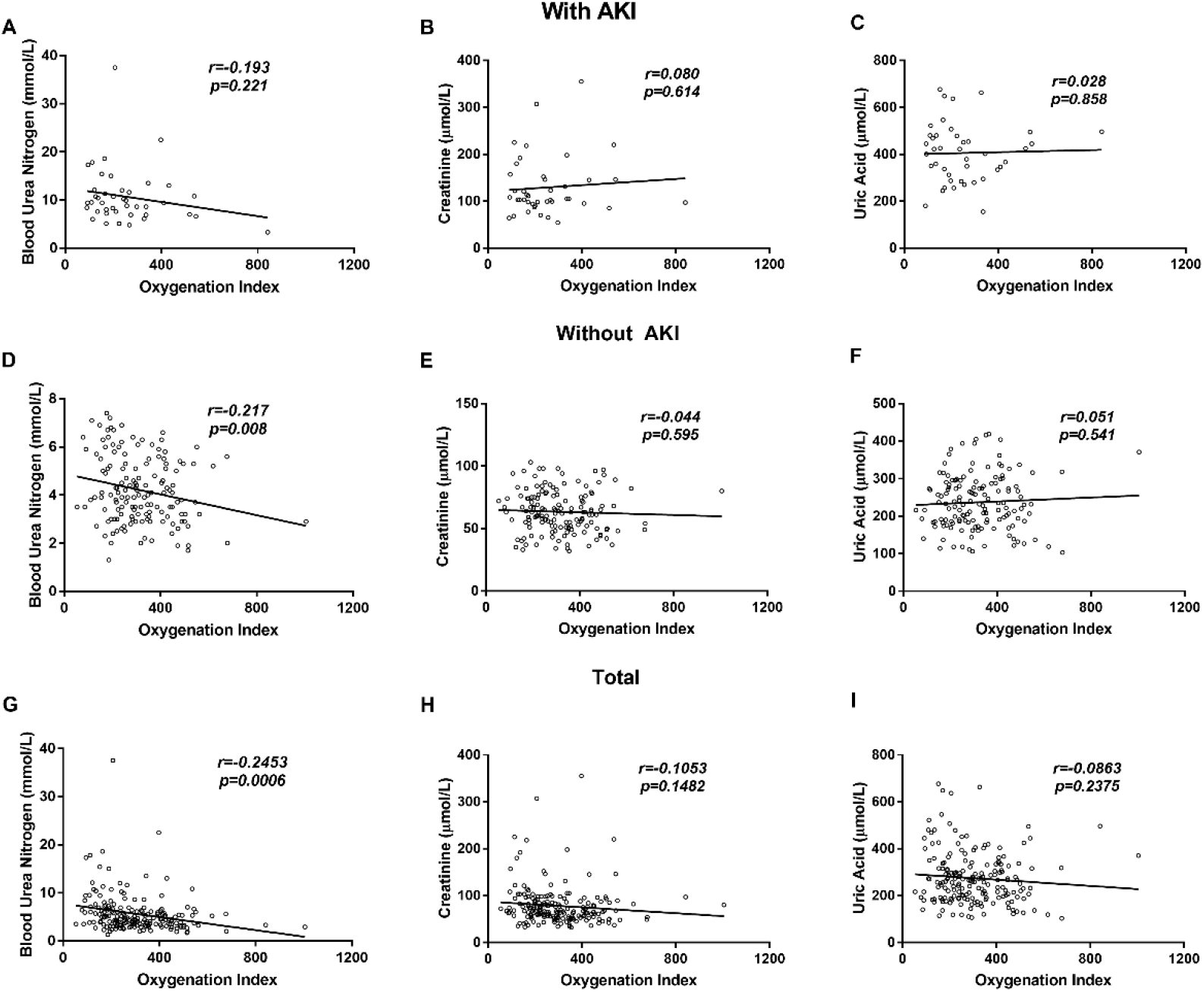
The association between oxygenation index and renal functional indices among COVID-19 patients. (A-C) Correlation between oxygenation index and renal functional indices was analyzed among COVID-19 patients with AKI. (A) Blood urea nitrogen; (B) Creatinine; (C) Uric acid. (D-F) Correlation between oxygenation index and renal functional indices was analyzed among COVID-19 patients without AKI. (D) Blood urea nitrogen; (E) Creatinine; (F) Uric acid. (G-I) Correlation between oxygenation index and renal functional indices was analyzed among all COVID-19 patients. (G) Blood urea nitrogen; (H) Creatinine; (I) Uric acid.

### 3.3 Male elderly COVID-19 patients with diabetes mellitus are more susceptible to AKI

The effects of different genders on renal functional indices were analyzed. As shown in Table 3, the levels of serum creatinine, urea nitrogen and uric acid were higher in males than those of females. The effects of ages on renal functional indices were then analyzed. As shown in Table 3, the levels of serum urea nitrogen were higher in patients over 70 years old than those from 50 to 69 years old and under 50 years old (Table 3). Moreover, the levels of serum urea nitrogen were higher in patients from 50 to 69 years old than those under 50 years old (Table 3). In addition, the levels of serum creatinine and uric acid were higher in patients over 70 years old than those under 70 years old (Table 3). The effects of comorbidity with either hypertension or diabetes on renal functional indices are shown in Table 4. The levels of serum creatinine, urea nitrogen and uric acid were higher in COVID-19 patients with diabetes than those without diabetes. No significant difference on serum creatinine, urea nitrogen and uric acid was observed between COVID-19 patients with hypertension and without hypertension (Table 4). Multivariable logistic regression was used to analyze risk factors of AKI among 355 COVID-19 patients. As shown in Table 5, the *OR* of males with AKI was 1.898 (95% *Cl*: 1.011, 3.562; *P*<0.05). The *OR* of older age with AKI was 1.976 (95% *Cl*: 1.264, 3.089; *P*=0.003). The *OR* of comorbidity with diabetes with AKI was 2.249 (95% *Cl*: 1.123, 4.505; *P*<0.05). No significant relationship was observed between comorbidity with hypertension and AKI among COVID-19 patients (Table 5).

**Table 3.** The effects of demographic characteristics and complications on renal function.

|  | Cases | BUN (mmol/L)<br>mean±SD | Cr (μmol/L)<br>mean±SD | UA (μmol/L)<br>mean±SD |
| --- | --- | --- | --- | --- |
| **Gender** |  |  |  |  |
| Female | 162 | 4.5±2.8 | 67.4±36.2 | 240.2±94.9 |
| Male | 193 | 5.3±3.6** | 76.5±32.9*** | 274.3±106.6*** |
| **Age** |  |  |  |  |
| <50 | 160 | 4.0±1.8 | 68.0±23.3 | 250.4±93.5 |
| 50-69 | 148 | 5.1±3.6*** | 72.0±39.5 | 250.3±103.8 |
| 70- | 47 | 7.5±4.4***, ### | 88.4±45.5**, ## | 313.8±114.1***, ### |
| **Hypertension** |  |  |  |  |
| No | 230 | 4.7±2.4 | 69.6±27.3 | 250.2±94.8 |
| Yes | 125 | 5.4±4.4 | 77.9±45.2 | 275.6±115.1 |
| **Diabetes** |  |  |  |  |
| No | 208 | 4.5±2.5 | 68.0±26.6 | 245.3±95.1 |
| Yes | 147 | 5.5±4.0** | 78.8±43.2* | 278.9±110.4** |

Note: Cases in Gender, compared with “Female”, \* $P<0.05$ , \*\* $P<0.01$ , \*\*\* $P<0.001$ .
Cases in Age, compared with “<50”, \* $P<0.05$ , \*\* $P<0.01$ , \*\*\* $P<0.001$ ; compared with “50-69”, # $P<0.05$ , ## $P<0.01$ , ### $P<0.001$ .
Cases in Hypertension, compared with “No”, \* $P<0.05$ , \*\* $P<0.01$ , \*\*\* $P<0.001$ .
Cases in Diabetes, compared with “No”, \* $P<0.05$ , \*\* $P<0.01$ , \*\*\* $P<0.001$ .

**Table 4.** Univariate analysis of AKI risk factors among COVID-19 patients.

|  | β | Wald | *P value* | OR (95% C.I.) |
| --- | --- | --- | --- | --- |
| **Male** | 0.641 | 3.979 | 0.046 | 1.898 (1.011, 3.562) |
| **Age** | 0.681 | 8.937 | 0.003 | 1.976 (1.264, 3.089) |
| **Hypertension** | 0.176 | 0.257 | 0.613 | 0.838 (0.424, 1.658) |
| **Diabetes** | 0.811 | 5.233 | 0.022 | 2.249 (1.123, 4.505) |

**Table 5.**
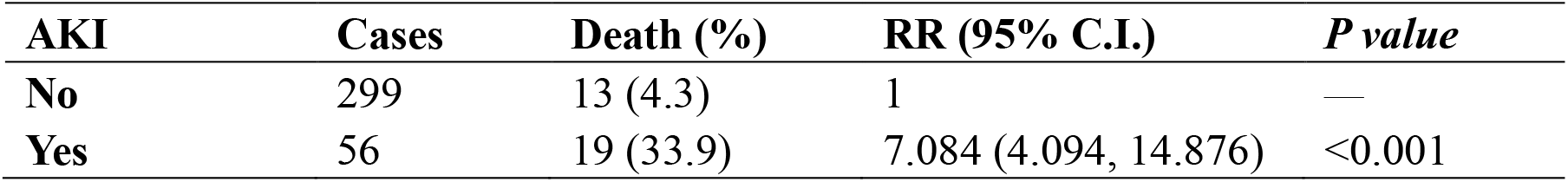
The association between AKI and death risk among COVID-19 patients.

| AKI | Cases | Death (%) | RR (95% C.I.) | *P value* |
| --- | --- | --- | --- | --- |
| **No** | 299 | 13 (4.3) | 1 | — |
| **Yes** | 56 | 19 (33.9) | 7.084 (4.094, 14.876) | <0.001 |

### 3.4 AKI at early stage elevates death risk of COVID-19 patients

The effects of AKI at the early stage on death risk were analyzed among COVID-19 patients. Among 56 COVID-19 patients with AKI, 33.9% were died on mean 10.9 day after hospitalization. The fatality rate was obviously higher among COVID-19 patients with AKI than those without renal injury (33.9 % vs 4.3%, *RR*=7.88, *P*<0.001).

## Discussion

The objective of the present study was to describe the clinical characteristics and analyze the influence factors of SARS-CoV-2-induced AKI among 355 COVID-19 patients in two hospitals from different regions. The major findings of the present study include: (1) AKI is more common in critical ill patients with COVID-19; (2) Male elderly COVID-19 patients with diabetes mellitus are more susceptible to AKI; (3) AKI at the early stage elevates death risk of COVID-19 patients.

Several studies indicated that COVID-19 patients were usually companied by multiple organ injuries, including myocardial dysfunction, hepatic injury and even acute renal failure ^13^. In the present study, we analyzed AKI at the early stage of COPID-19. AKI was determined by measuring biochemical indices including serum creatinine, urea nitrogen and uric acid. Our results showed that 15.8% COVID-19 patients were companied by AKI on admission. Off interest, the levels of serum creatinine, urea nitrogen and uric acid on admission were higher in critical ill patients than those of mild COVID-19 patients. Although some mild COVID-19 patients were with AKI on admission, AKI is more common in critical ill patients with COVID-19. These results suggest an association between AKI at the early stage and the severity of COVID-19 patients.

Increasing evidence demonstrated that elderly COVID-19 patients were more serious ^15,16^. In the present study, we analyzed the impact of gender and age on AKI at the early stage of COVID-19. We showed that the levels of serum creatinine, urea nitrogen and uric acid were higher in males than those of females. Moreover, the levels of serum creatinine, urea nitrogen and uric acid were higher in old patients over 70 years old than those of younger patients. Several reports showed that COVID-19 patients with comorbidities were more serious. Indeed, the present study found that more than half of COVID-19 patients were with either diabetes or hypertension. Thus, it is especially whether COVID-19 patients with either diabetes or hypertension are more susceptible to AKI. Unexpectedly, there was no difference on renal functional indices between COVID-19 patients with hypertension and without hypertension. Interestingly, urea nitrogen and uric acid were higher in COVID-19 patients with diabetes than those without diabetes. To exclude potential confounding factors, multivariable logistic regression was used to further analyze the impact of gender, age and comorbidities on AKI at the early stage of COVID-19. Our results indicated that male, older age and comorbidity with diabetes were major risk factors of AKI at the early stage of COVID-19 patients. Taken together, these results suggest that male elderly COVID-19 patients with diabetes mellitus are more susceptible to AKI.

An early study found that SARS patients with AKI were at an increased death risk^17^. In the present study, we analyzed the association of AKI with death risk of COVID-19 patients. Among 56 subjects with AKI on admission, about a third of patients died on mean 10.9 day after hospitalization. The fatality rate was obviously higher among COVID-19 patients with AKI than those without AKI. These results provide evidence that AKI at the early stage elevated death risk of COVID-19 patients.

The mechanism by which SARS-CoV-2 injection induces AKI is likely to be multifactorial. Several reports showed that most COVID-19 patients were companied with multiple organ damage ^9,14^. However, the present study found that there was no significant association between oxygenation index and all measured renal functional indices among COVID-19 patients with AKI, suggesting that SARS-CoV-2-induced AKI is independent of respiratory failure. Two early reports showed that ACE2, as a receptor for SARS-CoV-2, was highly expressed in renal tubular epithelium ^18,19^. Another early study found that SARS viral particles were detected in the epithelium of the renal distal tubules ^20^. Therefore, the present study does not exclude that SARS-CoV-2 induces AKI through directly infecting renal epithelium. Further experiments are required to explore whether renal tubular epithelium is another target of SARS-CoV-2 injection.

In summary, the present study described the clinical and laboratory characteristics of AKI among 355 COVID-19 patients in two hospitals from different regions. Our results showed that AKI was more common in critical ill patients with COVID-19. Moreover, male elderly COVID-19 patients with diabetes mellitus were more susceptible to AKI. We found that AKI at the early stage elevated death risk of COVID-19 patients. We provide evidence that the development of AKI at the early stage may be a potential negative prognostic indicator for survival of COVID-19 patients. Therefore, the improvement of renal function is beneficial for elevating the survival rate of COVID-19 patients especially critical ill patients.

## Data Availability

All data used to support the findings of this study are available from the corresponding author upon request.

## AUTHOR CONTRIBUTIONS

D.X.X., H.Z. designed research; S.X., L.F., J.F., H.X.X., Y.X., Z.X.T, M.D.L, F.F.L., H.Y.L., L.Z. and Y.L. conducted research; S.X., L.F. analyzed data; D.X.X. and S.X. wrote the paper; D.X.X. and L.F. had primary responsibility for final content. All authors read and approved the final manuscript.

## ACKNOWLEDGMENTS

We thank all members of respiratory and critical care medicine in the Second Affiliated Hospital of Anhui Medical University, Union Hospital of Huazhong University of Science and Technology and The Second People’s Hospital of Fuyang City for recruiting participators.

